# Risk factors for long-term decline in kidney function amongst Malawian adults living in rural Karonga: protocol for a prospective cohort study

**DOI:** 10.1101/2025.08.01.25332688

**Authors:** Charlotte M Snead, Chimwemwe Mkandawire, Paul Kambiya, Shekinah Munthali-Mkandawire, Fredrick Kalobekamo, Albert Dube, Thandile Nkosi-Gondwe, Baltazar Bananga Mtenga, Dominic Nzundah, Lenford Kwamkwanya, Desire Bellings, Wisdom Nakanga, June Fabian, Robert Kalyesubula, Chimota Phiri, Felix Limbani, Dominic M Taylor, Amelia C Crampin, Henry C Mwandumba, Alison J Price

**Affiliations:** Liverpool School of Tropical Medicine, Pembroke Place, Liverpool, L3 5QA, United Kingdom; Malawi Epidemiology and Intervention Research Unit, PO Box 46, Chilumba, Malawi; Malawi Liverpool Wellcome Research Programme, Queen Elizabeth Central Hospital Campus, Chipatala Avenue, PO Box 30096 Chichiri, Blantyre 3, Malawi; London School of Hygiene and Tropical Medicine, Keppel Street, London, WC1E 7HT, United Kingdom; University of Glasgow, University Avenue, Glasgow, G12 8QQ, United Kingdom; Deanery of Clinical Sciences, College of Medicine and Veterinary Sciences, University of Edinburgh, 49 Little France Crescent, The Chancellor’s Building, Edinburgh, EH16 4SB, United Kingdom; Wits Donald Gordon Medical Research Institute, Faculty of Health Sciences, University of Witwatersrand, 7 York Rd, Parktown, Johannesburg, South Africa; Makerere University, 7062 University Road, Kampala, Uganda; Kamuzu University of Health Sciences, Private Bag 360, Chichiri, Blantyre 3, Malawi; Population Health Sciences, Bristol Medical School, University of Bristol, Canynge Hall, 39 Whatley Road, Bristol, BS8 2PS, United Kingdom; Richard Bright Renal Service, North Bristol NHS Trust, Southmead Road, Bristol, BS10 5NB, United Kingdom

## Abstract

**Background:** The global burden of chronic kidney disease (CKD) is rising, disproportionately impacting on low- and middle-income countries. In African populations, use of serum creatinine to estimate glomerular filtration rate (GFR) significantly underestimates CKD prevalence, contributing to its under-recognition as a health problem. Serum cystatin C provides more accurate estimates of Iohexol measured GFR than creatinine. Early diagnosis and treatment of CKD, targeted towards high-risk individuals, is essential to reduce premature morbidity and mortality. However, little is known about risk factors for CKD development and progression in Africa owing to limited longitudinal data. This study aims to determine risk factors for progressive kidney function decline among adults living in rural, northern Malawi.

**Methods:** This protocol describes an ongoing prospective study being conducted in a general population cohort in rural Karonga, Malawi. We are recruiting adults aged 18 years and over who participated in two previous population-based surveys of long-term health conditions, over five years apart. New household-level data is being collected on CKD risk factors, alongside blood and urine samples. Cystatin C and creatinine will be tested on individual-level paired, stored serum samples collected at three longitudinal time points. Urine will undergo dipstick urinalysis, microscopy and testing for albumin and creatinine to quantify proteinuria. The primary outcome will be sustained 25% reduction in estimated GFR (eGFR) from baseline and change in eGFR category, determined using serum cystatin C. Multivariable logistic regression will be used to determine effect size estimates of key risk factors for kidney function decline.

**Discussion:** This study will provide important data on risk factors for eGFR decline and CKD progression amongst Malawian adults. The findings will inform future research into important context-specific risk factors, and could directly inform future health policies in Malawi for targeting CKD screening, prevention and treatment strategies to the highest risk patient groups.

## Introduction

The global burden of chronic kidney disease (CKD) and associated mortality is rising(1), with CKD predicted to be the fifth leading cause of life years lost by 2040.(2) At the 78^th^ World Health Assembly in May 2025, the World Health Organization (WHO) adopted a long-awaited resolution recognising CKD as a global health priority and committing to promotion of kidney health through prevention and control of kidney disease.(3) The burden is disproportionately impacting on low- and middle-income countries (LMIC), where over 77% of people living with CKD reside, the majority undiagnosed.(4) Set against this, stark disparities in access to appropriate diagnostic tests, prevention and treatments remain.(5) Only 7% of individuals receiving kidney replacement therapy (KRT) live in LMIC(6) and there are huge workforce shortages for delivery of kidney care. Compared to high-income countries, nephrologist densities in low-income counties and lower-middle income countries are more than 80 times and 10 times lower, respectively.(7) Notably, Malawi has only two adult nephrologists for a population of over 21 million people.

In most of Africa, the prevalence of CKD is thought to be between 13 and 15%.(8–10) However, accurate estimates of burden are hampered by limited access to essential diagnostic blood and urine tests(11, 12), heterogeneity of methods for measuring CKD prevalence across studies(10), and uncertainty about the most appropriate methods for estimating kidney function in African settings.(13) Importantly, the most readily available creatinine-based equations for estimating glomerular filtration rate (eGFR_creat_) have been shown to perform extremely poorly in African populations, significantly underestimating the overall burden of CKD when compared to gold-standard measured GFR (mGFR) determined by Iohexol plasma clearance.(13) This has further contributed to under-recognition of CKD as a public health problem in many African countries.(13) In Malawi, population prevalence of eGFR less than 60ml/min/1.73m^2^ determined using the internationally recommended creatinine-based CKD-EPI (2021) equation(14) is only 0.49% (95% CI 0.31 – 0.73) and using the CKD-EPI 2009 equation(15) without adjustment for African-American ethnicity, 1.25% (95% CI 0.95 – 1.61), compared to 11.91% (95% CI 10.13 – 13.69) when using imputed GFR modelled on Iohexol mGFR.(13) Estimating GFR using serum cystatin C (eGFR_cysc_) more closely mirrors mGFR in African populations(13), but this assay is significantly more expensive than serum creatinine and is not currently routinely clinically available in African settings.

Given the limited available resources in many LMIC for treating advanced kidney disease, early detection, prevention and treatment, targeted towards those at highest risk, is essential. Early initiation of disease-modifying treatments to slow CKD progression, reduce risk of kidney failure and mitigate CKD-related complications will reduce premature morbidity and mortality, including the significantly heightened risk of CKD-attributable cardiovascular morbidity and mortality.(5, 16–21)

CKD fits the WHO criteria for screening that has public health benefit, given its asymptomatic nature and the existence of available treatments, however evidence for cost-effectiveness of mass population screening is lacking(22–25). Instead, screening in high-risk groups, including individuals with diabetes and hypertension, is recommended, provided treatment can be provided.(26–29) However, in LMIC, including those in Africa, the rising prevalence of NCDs intersects with ongoing high burdens of infectious diseases, poverty, malnutrition, maternal health challenges, environmental and climate factors. As a result, the potential causes of CKD are diverse, and the groups of individuals at highest risk for CKD progression may therefore differ from those in high income settings.(30–34) Indeed, several cross-sectional studies conducted across East and Southern Africa have suggested that only one-fifth to two-thirds of CKD appears to be associated with traditional risk factors.(35–38) As such, it remains unclear in many African settings which individuals should be prioritised for CKD screening and prevention programs, above and beyond those with hypertension, diabetes and proteinuria.(22)

Improved knowledge on the individuals at highest risk for rapid eGFR decline, CKD development, progression and future kidney failure would help target resources to those at the highest risk of adverse CKD outcomes, and would also contribute to future development of context-specific clinical risk prediction tools, (not yet validated in African settings(39, 40)) to aid identification of high-risk patients, risk-stratification and prognostication. Understanding this requires longitudinal studies of kidney function trajectories, however very few studies of this nature have been conducted in African settings. Those that exist have generally been conducted in hospital settings(41–44), amongst specific patient groups(42, 43, 45–49), and/or over relatively short follow-up periods.(41, 42, 44, 49) Furthermore, all have assessed eGFR trajectories using creatinine-based eGFR measures (eGFR_creat_), and not using cystatin C (eGFR_cysc_), now known to be a much more accurate predictor of kidney function in African populations.

### Aim and objectives

The aim of this prospective cohort study is to determine key risk factors for progressive decline in eGFR amongst adults participating in a general population cohort in rural northern Malawi. The primary objective is to determine predictors for progressive decline in eGFR_cysc._ A secondary objective is to determine risk factors for decline in eGFR_creat_ and development of CKD.

## Methods

### Study design and setting

The “Impso” (“kidney” in Chichewa, a local language) study is a longitudinal, prospective cohort study set within Karonga Health and Demographic Surveillance System (HDSS), an open, population-based cohort established in 2002 and covering an area of 135km^2^ of rural subsistence farming and fishing communities in northern Malawi(50) managed by the Malawi Epidemiology and Intervention Research Unit (MEIRU) **[Figure 1].**

**Figure 1.**
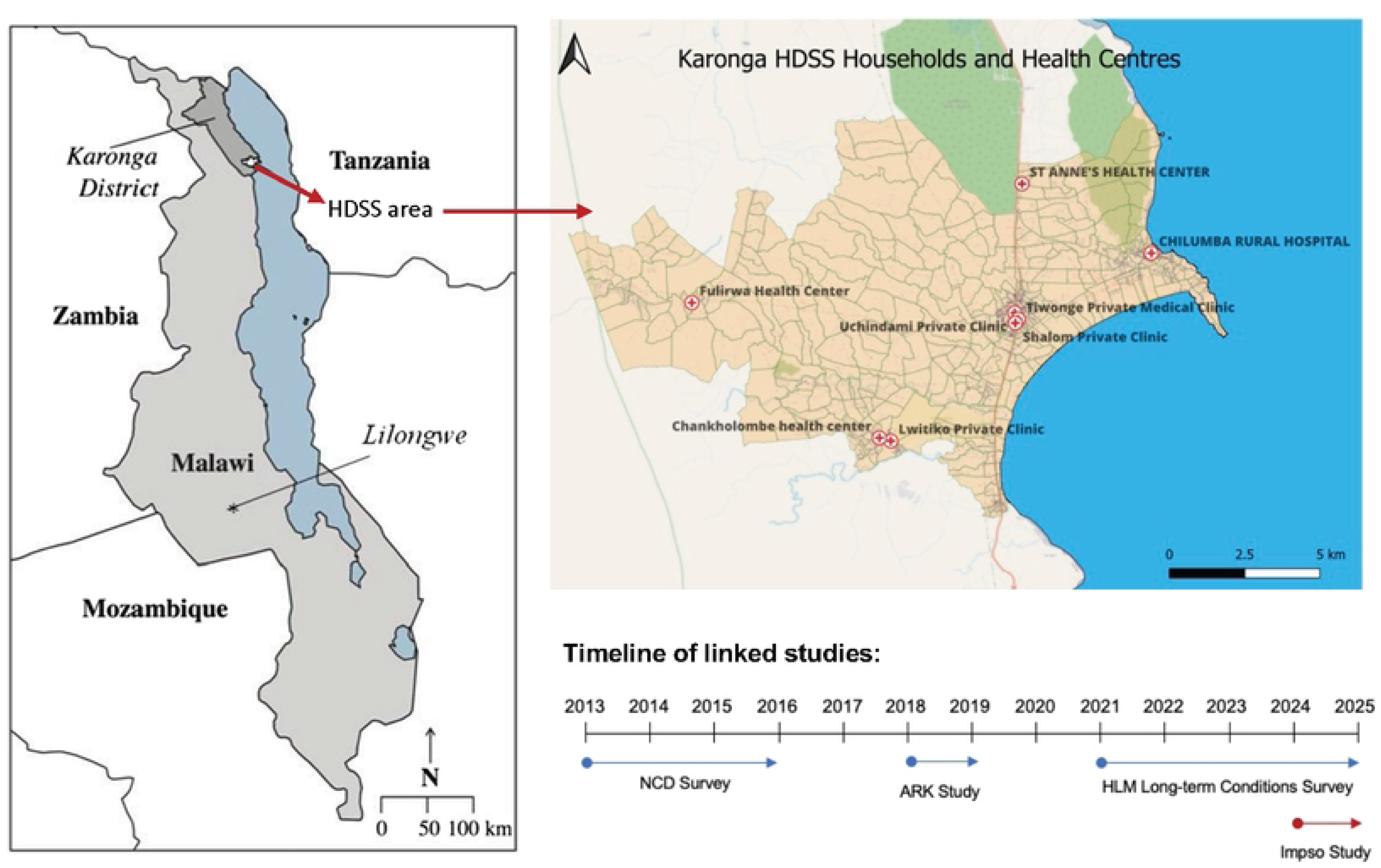
The Karonga HDSS and linked studies.

The HDSS includes a population of over 53,000 (data to early 2025) and is served by four health facilities (rural hospitals and health centres)(50). The nearest secondary referral hospital is located 80km to the north in Karonga boma. The HDSS undergoes continual population surveillance of births and deaths, with annual census of inward- and outward migration and collection of detailed sociodemographic data. All deaths undergo verbal autopsy (VA) with physician-assigned cause of death following a medical interview and case review using the standardised WHO VA tool.(50)

The Karonga HDSS, alongside a more recently established peri-urban HDSS in the outskirts of Lilongwe, is the site of two comprehensive population-based surveys of long-term health conditions (LTCs): a non-communicable disease (NCD) survey of cardiometabolic conditions conducted 2013-16(51, 52); and the currently ongoing Healthy Lives Malawi (HLM) LTC survey; 2021-25(53), which aims to re-survey prevalence and assess progression of cardiometabolic conditions, in addition to newly determining prevalence of additional LTC and more detailed risk factor data. Both studies contribute to a biobank.

The currently described study capitalises on these two larger studies **[Figure 1]** and additionally leverages data from a large multinational study conducted in 2018-19 by the African Research in Kidney Disease (ARK) network to investigate methods for measuring kidney function in African populations(13), which included 510 adults residing in the Karonga HDSS. Available data from all three linked studies includes household data (socioeconomic status; geolocators), interview data (demographics; lifestyle factors; clinical history, including prior diagnosis, screening and medications for chronic conditions), examination and physical measures (anthropometry; blood pressure; hand grip strength; peripheral arterial measures) and biological samples (serum, plasma and whole blood samples stored at -80 Celsius and other biological material). Baseline cystatin C and creatinine have already been tested on stored serum samples collected between 2013 and 2019(13, 38) for 2792 and 4637 adults respectively.

### Participants

This study is recruiting adults living within the Karonga HDSS, who meet the following eligibility criteria **[Figure 2]**:

- Alive, self-defined as resident within the Karonga HDSS^1^, and provided consent to ongoing participation in the HDSS and sampling for further studies.
- Participated in both the NCD (2013-16) and HLM-LTC (2021-25) population surveys.
- Aged ≥ 18 years at the time of participation in the NCD (2013-16) survey.
- Availability of baseline cystatin C tested on a stored historical serum sample.
- Baseline eGFR_cysc_ <90 ml/min/1.73m^2^.

**Figure 2.**
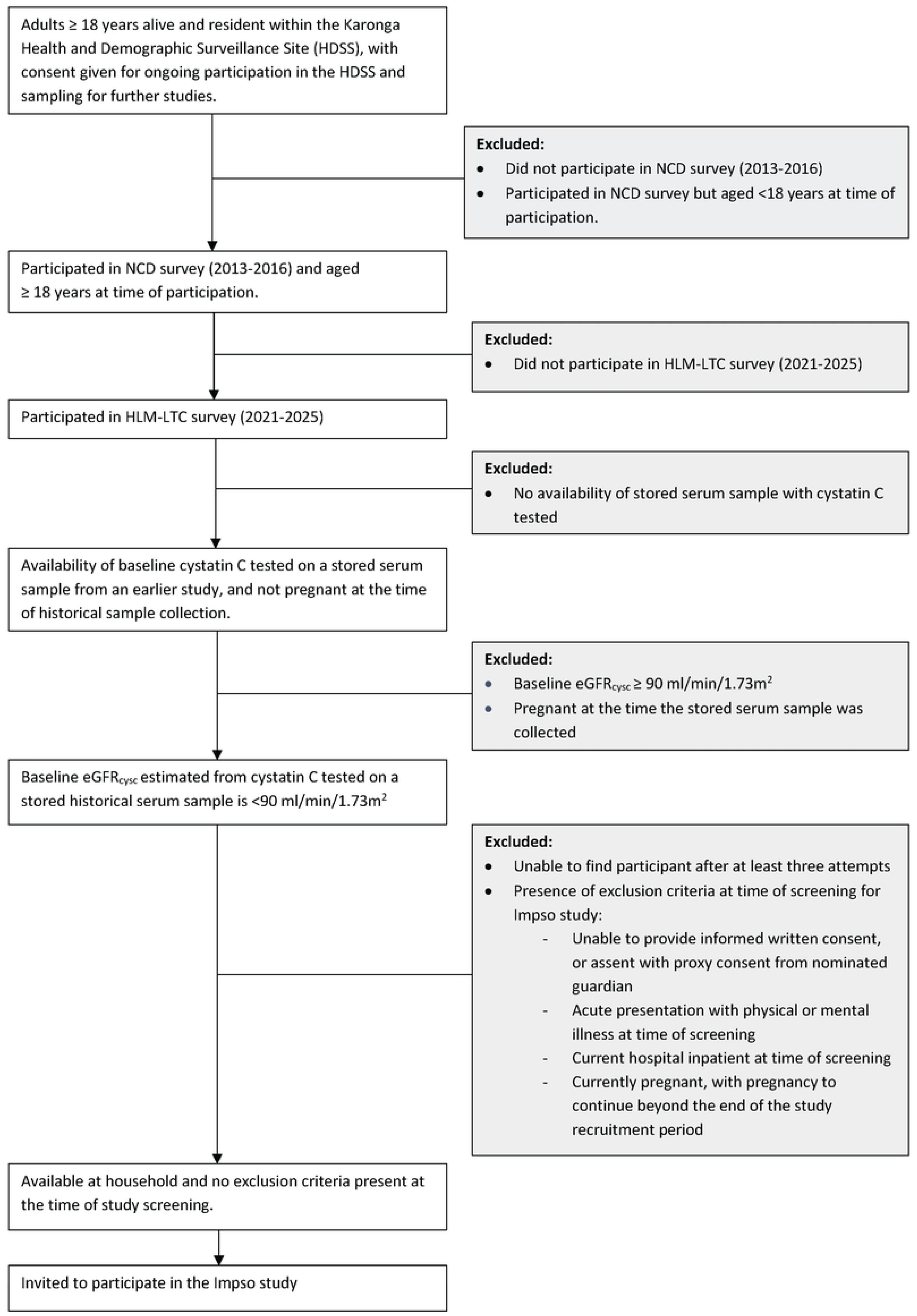
Participant selection process

Exclusion criteria include:

- Unable to provide informed written consent, or assent with proxy informed written consent from a nominated guardian.
- Acute presentation of physical or mental illness or rapidly declining poor health at the time of attempted recruitment, which might be exacerbated by participation in the study.
- Currently a hospital inpatient at time of follow-up (at time of screening for participation in Impso study)
- Pregnant at the time of baseline sample collection for cystatin C testing, and/or when approached for recruitment into the current study, with pregnancy to continue beyond the end of the study recruitment period.

### Recruitment and consent

Individuals potentially meeting the inclusion criteria are identified by screening the MEIRU database. For this purpose, the MEIRU-authorised study team members were granted access to linked datasets (secured in encrypted servers, only accessible by authorised personnel through provided credentials) on 08/08/2023 for the purposes of screening, study visits and results dissemination. All participants in the datasets have given consent for use of their data in future linked studies. Final datasets are stripped of personal identifying data. No study team members have access to identifying **after** data collection and results dissemination are complete. All study team members undergo training in good clinical practice in research.

Potentially eligible participants are approached at their household to invite participation in the study, a minimum of three months after their participation in the HLM LTC survey. Up to three attempts are made to visit each potentially eligible participant, including at different times of day, to facilitate recruitment of adults of working age who may be less likely to be at home and thereby minimise selection bias. After checking for exclusion criteria, individuals who accept receive information about the study in the form of a participant information sheet, provided in their preferred language (Chitumbuka, Chichewa or English). The information sheet is read aloud to the potential participant by a member of the study team and the individual is given the opportunity to ask questions.

Individuals who wish to take part proceed to provide informed, written consent. Individuals who are unable to read or write provide a thumbprint instead of a signature. In this event, an impartial witness is present throughout the information-giving and consent process, and also signs the informed consent form. For individuals who lack capacity to provide informed consent, but are able to provide assent, proxy consent is sought from the nominated guardian. Informed written consent is obtained for all planned study activities, including for long-term storage of study data and biological samples.

### Household-level data collection

Data collection takes place over the course of two to three household visits **[Figure 3].**

**Figure 3.**
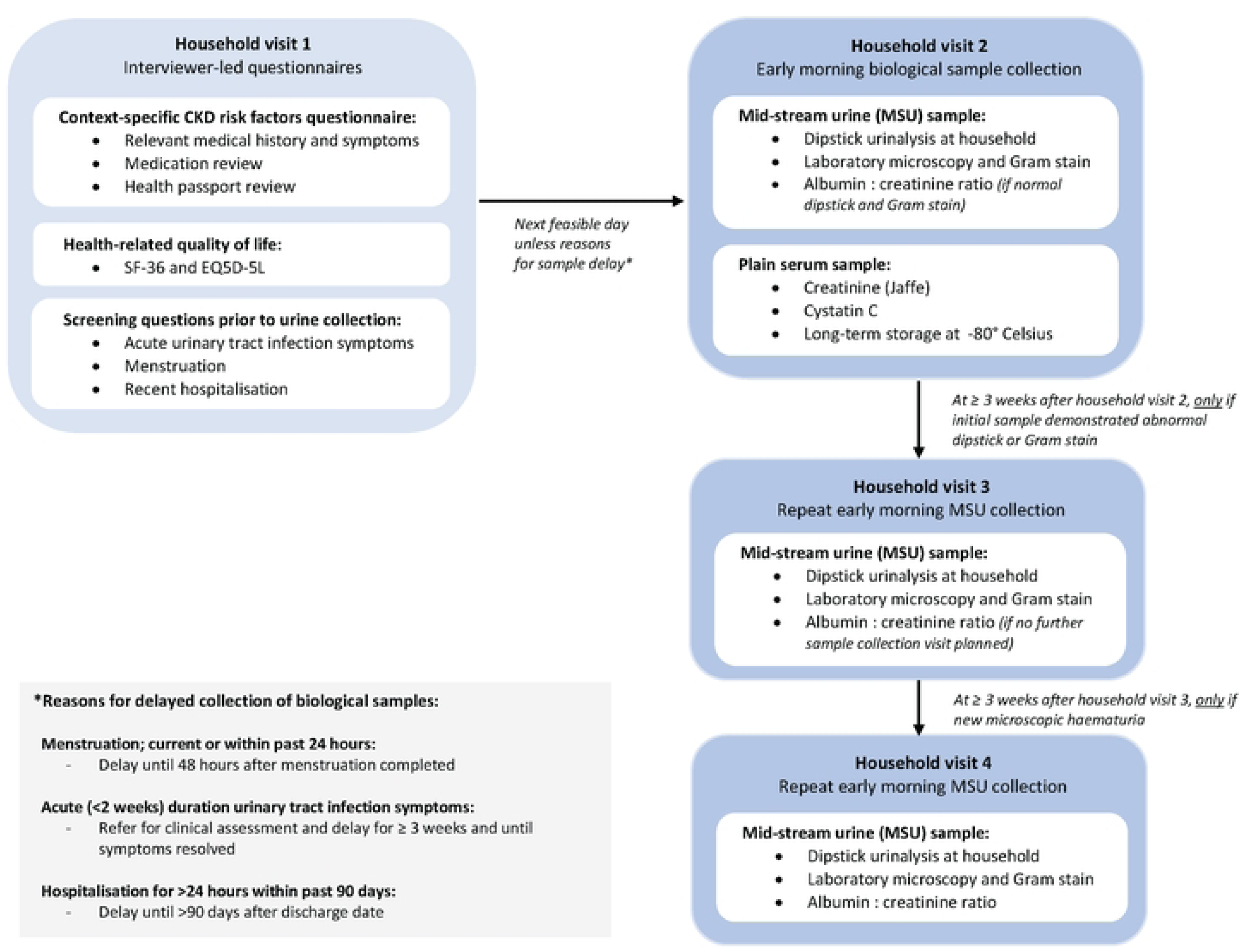
Impso study data collection activities.

#### Interviewer-led questionnaires

The first household visit involves delivery of several questionnaires, covering; (1) context-specific risk factors for CKD, including detailed questions on medical history, symptoms and medication use, guided by *a priori* hypotheses on exposures of importance in this setting(31, 38), and supported by review of the participant’s health passport if available; (2) health-related quality of life (HrQOL), using the RAND 36-Item Short Form Survey Instrument (SF-36), and the EQ-5D-5L, both translated into Chitumbuka and Chichewa, with the SF-36 adapted where appropriate for relevance to the Malawi setting(54), and use of the EQ-5D-5L translations (already validated in Chichewa)(55) approved by EuroQol; and (3) screening questions to determine optimal timing for early morning biological sample collection. Sample collection is delayed in the event of urinary tract infection symptoms, current or recent (within past 24 hours) menstruation, and hospital admission >24 hours duration within the preceding 90 days [**Figure 3].**

HIV testing and counselling (HTC) using point-of-care Determine^TM^ rapid test kits is offered to participants who have never been tested, are unsure about their HIV status, or whose most recent test with a non-reactive result was conducted over six months previously (ascertained from both self-report and health passport documentation). Study staff performing HTC have undergone formal training and certification though Malawi Ministry of Health HTC training providers. Any participants with a reactive result on the Determine^TM^ rapid HIV test are referred to the HIV clinic at Chilumba Rural Hospital for repeat counselling and full testing using the three-test diagnostic algorithm, following Ministry of Health guidelines.(56)

All questionnaire data is captured electronically on Android tablets using the Open Data Kit (ODK) platform(57, 58). Before completing the first household visit, the study staff ensure that clear instructions have been given to participants in preparation for mid-stream urine (MSU) sample collection using a standardised information sheet provided in Chitumbuka, Chichewa or English **[Supplementary file S1].**

#### Biological sample collection

Collection of biological samples (MSU and single venepuncture serum sample) takes place at a second, early morning household visit, the day after the interview date, or else on the first alternative convenient date for the participant. Immediately prior to sample collection, study staff repeat screening questions for menstruation and UTI symptoms, and check that the MSU collection instructions have been followed. MSU samples are collected in sterile containers, contained with sealed sterile packaging until the time of collection, and are inspected by the study team prior to dipstick urinalysis testing. Samples collected incorrectly, that are clearly contaminated, leaking, or that are inadequate (< 20 ml) in volume are discarded without testing and arrangements are made for re-collection.

Dipstick urinalysis is being conducted on acceptable MSU samples at the household using Siemens Multistix*^®^* 10 SG urine dipsticks following a strict standard operating procedure, including use of a sterile Pasteur pipette to apply urine to dipstick test pads, and use of an electronic timer to ensure dipstick results for each reagent are read at the times specified by manufacturer. Urine dipstick results are captured electronically at the household using ODK and are photographed at two minutes for quality control purposes. Any abnormalities on dipstick urinalysis triggers a third household visit in ≥ three weeks’ time for collection and testing of a repeat (second) MSU sample. In the event of microscopic haematuria newly present on a second sample, a third and final MSU collection visit takes place **[Figure 3].** Venepuncture serum samples will be collected in 10ml plain serum tubes using vacutainer needles.

Samples are placed upright in cool boxes maintained at 2 – 8 °C immediately after collection and urinalysis, and are transported back to the MEIRU laboratory within four hours of sample collection. Within this four-hour window, urine samples spend a maximum of two hours at ambient temperature prior to the cold storage.

### Laboratory methods

The condition of all biological specimens is inspected on arrival at the laboratory; samples that are leaking, contaminated or inadequate are rejected and arrangements are made for recollection.

Urine microscopy involves both wet mount examination and Gram stain of urine sediment. After mixing by inversion, 10 ml from each mid-stream urine sample is centrifuged at 720 g for five minutes immediately after reaching the laboratory. After separating from the supernatant, urine sediment is emulsified and the sediment examined by light microscopy at both low power field (LPF, x10 objective) and high power field (HPF, x40 objective), the latter for identification and quantification of cells (erythrocytes, leucocytes, epithelial cells), casts, crystals and Schistosoma ova. Cell counts are reported per HPF, and Schistosoma ova per 10 ml of urine.

Prior to Gram stain, urine sediment is mounted on the slide and gently heat fixed for five minutes. Gram stain is then performed by sequentially flooding slides, first with 0.5% crystal violet (left for 60 seconds), rinsed off with water, then with Gram’s iodine (left for 60 seconds), washed off with acetone decolouriser before rinsing with water, and finally with safranin (left for 30 seconds), again rinsed off with water. After blotting dry, slides are examined using x100 objective lens with emersion oil. Participants with organisms identified on Gram stain of the baseline urine sample receive a follow-up visit for repeat sample collection, with reiteration of MSU collection instructions. Urine culture is not being performed owing to budget constraints of the study.

The remaining uncentrifuged portion of the urine sample for each participant is mixed by inversion and used to extract three 1ml aliquots using 2ml cryotubes, which are stored at -20°C temporarily, then transferred to the -80°C freezer within 12 hours of sample collection for short-term storage until the time of urine chemistry analysis, which will take place after participant recruitment is complete.

Blood samples collected in plain tubes are centrifuged within two hours of reaching the laboratory by spinning for ten minutes at 1620g in a swing bucket centrifuge. The separated serum is used to make five aliquots of 0.5ml using 2ml cryotubes, stored at -20°C temporarily, then transferred to -80°C freezer within 12 hours of sample collection for long-term storage.

Prior to chemistry testing, stored serum and urine samples will be removed from the -80°C freezer and allowed to thaw for 30 minutes to 1 hour. After completing thawing, samples will be vortexed and put in sample cups ready for testing. All chemistry testing will use Beckman Coulter AU480 chemistry analysers. Serum samples from this study, and individual level paired serum samples from the HLM-LTC survey, will be tested for serum cystatin C (immunoturbidimetric method, standardised using ERM-DA471/IFCC reference material) and creatinine (kinetic compensated Jaffe method, method A, standardised to an isotope-dilution mass spectroscopy [IDMS] assay). For cystatin C testing, serum will be mixed with Gentian Cystatin C reagent containing cystatin C immunoparticles. Cystatin C from the serum and anti-cystatin C from the immunoparticles form aggregates. The complex particles created absorb light, and by turbidimetry the absorption is related to cystatin C concentration via interpolation on an established standard calibration curve. The AU480 platforms automatically calculate the results. The machine will be calibrated using Gentian Cystatin C Calibrator which is standardised against the international calibrator standard ERM-DA471/IFCC. A six-point standard curve will be established using standards one to six with assigned values given on the analytical value sheet provided with the calibrator. In the kinetic compensated Jaffe method, creatinine will form a coloured compound with picric acid in alkaline medium. The rate of change in absorbance at 520/800nm is proportional to the creatinine concentration in the sample. Interference from protein will be mathematically corrected by subtracting 18 μmol/L from each test result. The Beckman Coulter AU480 analyser will automatically compute the creatinine concentration of each sample. Serum calibrator creatinine value for method A is traceable to the Isotope Dilution Mass Spectroscopy (IDMS) method via National Institute of Standards and Technology (NIST) Standard Reference Material (SRM) 967.

Urine will be tested for albumin (turbidimetric method) and creatinine (kinetic compensated Jaffe method). In the turbidimetric method for urine albumin testing, anti-human serum albumin antibodies combine with albumin from the sample to form immune complexes that scatter light in proportion to their size, shape and concentration. The absorbance of these aggregates is proportional to the albumin concentration in the sample. Change in absorbance is measured at 380nm with subtraction of a reference wavelength at 800nm. The urine calibrator creatinine value is traceable to the IDMS method. Urine albumin calibrator values are traceable to the International Federation of Clinical Chemistry Certified Reference Material CRM470.

Urine samples will be discarded after validation of results; but serum samples will be stored long-term to allow additional testing in future studies.

#### Laboratory quality control processes

Prior to microscopic examination of urine (for both Gram staining and wet mount examination of urine sediment) the performance of the objective lenses is verified using standardised positive and negative control slides. Quality control of the microscopy procedures is maintained by independent verification of findings by two qualified technologists. During serum cystatin C and creatinine testing, low and high concentration controls will be tested each day before any samples are measured to validate the calibration curve. The controls have an assigned value range that will be met before measuring samples; these are given in the Analytical Value sheet included with the Gentian Cystatin C and creatinine control kits. During urine albumin and creatinine testing, low and high concentration controls of Biorad Liquichek assayed quality control material will be tested a minimum of once a day on each day of testing.

### Pilot phase

The data collection tools and field activities strategy were piloted both internally and externally to understand and address in a timely manner any issues associated with the workflow, logistics and time required, for the information giving, consent, medical interview, and biological sample collection processes. During this pilot phase, the questionnaire and field procedures were reviewed and revised based on feedback raised. This included improvements to some questionnaire questions, answer options and translations to improve understandability and cultural acceptability, in addition to revision of working procedures for more efficient collection of interview data and biological samples.

### Timelines

Recruitment for the pilot phase took place between 14 February and 1 March 2024. Recruitment for the main study commenced on 5 March 2024 and is ongoing, anticipated to complete by 30 September 2025. Data collection is anticipated to complete by 30 November 2025, and results are anticipated by March 2026.

### Analysis

#### Sample size

The target sample size is 1000 participants. A sample of n=1000 participants with eGFR_cysc_ available at baseline will give ≥80% power to detect odds ratio (OR) of between 1.66 and 4.38 for the effect of risk factors on eGFR decline. This is assuming that; (1) the risk factors of interest increase probability and rate of eGFR decline; (2) risk factor prevalences range between 5% and 30% (38, 51); and (3) eGFR decline outcome measures occur in 5 to 15% of individuals unexposed to the risk factors of interest.

#### Outcome measures

The study outcome measures are guided by the Kidney Disease Improving Global Outcomes (KDIGO) as well as the National Kidney Foundation and Food and Drug Administration guidance on surrogate end points for CKD progression.(59, 60) In each of the below definitions, for the purposes of this study, the term “sustained” means that the outcome is present on testing of two serum samples, collected a minimum of four weeks apart (but aiming for over 90 days apart; one sample collected in the 2021-25 HLM LTC survey, and other in the currently described 2024-25 Impso study), when compared to results from testing of the baseline sample (2013-19) sample. “Non-sustained” means that the outcome has been reached for the first time at the point of the most recent (Impso study) sample collection.

The primary outcome measure will be a sustained 25% reduction in eGFR_cysc_ from baseline and change in eGFR_cysc_ category.

The secondary outcome measures will be:

1. Non-sustained 25% reduction in eGFR_cysc_ from baseline and change in eGFR_cysc_ category
2. (a) Sustained and (b) non-sustained 25% reduction in eGFR_creat_ from baseline <u>and</u> change in eGFR_creat_ category
3. (a) Sustained and (b) non-sustained decline in (a) eGFR_cysc_ and (b) eGFR_creat_ by:

- 57% (equates to doubling of serum creatinine)(60)
- 40%
- 30%
4. Estimated annual decline* in (a) eGFR_cysc_ and (b) eGFR_creat_ of:

- -5ml/min/1.73m^2^/year
- -3ml/min/1.73m^2^/year
- *\*(Data from other settings suggests that age-related GFR decline in healthy adults without hypertension is no more than −1.07 mL/min/1.73m^2^/year)*(*61–63*)
5. Incident CKD, defined using eGFR criteria as:

- Sustained and (b) non-sustained new eGFR <60ml/min/1.73m^2^ (not present at baseline)
6. Kidney failure, defined as:

- Sustained and (b) non-sustained eGFR_cysc_ <15ml/min/1.73m^2^; or
- Sustained and (b) non-sustained eGFR_creat_ <15ml/min/1.73m^2^; or
- Commencement of maintenance KRT (≥4 weeks duration)^2^

#### Statistical analysis

Logistic regression (univariate and multivariate models) will be used to determine effect size estimates (odds ratios) of key covariates (risk factors) for kidney function decline (primary and secondary outcomes) to interrogate how much of kidney disease progression is explained by potential novel risk factors of interest, such as self-reported history of severe malaria or sepsis, HIV infection, previous TB or schistosomiasis infection, use of non-steroidal anti-inflammatory medications, use of traditional medicines, use of antibiotics, pregnancy-related complications and occupational exposures. Adjustment will be made for traditional risk factors (hypertension, diabetes, smoking and cardiovascular disease), established confounders (age, sex, body mass index, socioeconomic status) and for baseline eGFR. Potential interaction between sex and the effect of risk factors on outcome measures will be examined, as there is evidence from other settings that sex may modulate the relationship with renal function decline, in particular cardiometabolic risk factors.(64) Heterogeneity by age in the associations with outcome measures will also be investigated. In addition linear mixed effects models will be used to determine eGFR trajectories over time and estimate differences in eGFR slopes according to baseline risk factors.(65)

#### eGFR equations

All eGFR_cysc_ values will be calculated using the CKD-EPI 2012 equation.(66) eGFR_creat_ will be calculated using the CKD-EPI (2009) equation(15) without adjustment for African American ethnicity, as this has been demonstrated to have slightly higher accuracy and lower relative bias than the newer CKD-EPI (2021) equation when compared to Iohexol mGFR in African populations, including in adults from rural Karonga.(13) However sensitivity analyses for will also be conducted using the CKD-EPI (2021) creatinine equation (which has no ethnicity coefficient), the CKD-EPI (2021) combined creatinine-cystatin C equation and the European Kidney Function Consortium (EFKC) equations for both creatinine and cystatin C.(14, 67–69)

### Ethics, Governance And Regulations

#### Ethical approval

This study was approved by the National Health Sciences Research Committee (NHSRC) in Malawi (protocol #23/04/4049) and by the Liverpool School of Tropical Medicine, UK (protocol #23-019).

#### Feedback of results and clinical referral

Urine dipstick results will be communicated to study participants immediately after testing and will be recorded in health passports. Participants with abnormal baseline urine results requiring follow-up urine collection (see ‘Biological Sample Collection’), will receive results of the laboratory urine microscopy at the follow-up visit.

Baseline eGFR results are communicated to participants at the point of recruitment, accompanied by provision of general CKD lifestyle advice using a standardised information sheet. Repeat kidney function results (eGFR, uACR) obtained by testing of serum and urine samples collected in this study, and on paired LTC samples, as well as remaining normal urine test results, will be communicated once recruitment and testing is complete. **Supplementary file S2** outlines the indications for clinical referral and assessment. Clinical assessment will take place at an NCD clinic at Chilumba Rural Hospital, which was established by MEIRU to support the wider HLM LTC survey. Any onward referrals required to secondary or tertiary facilities will take place via existing referral pathways.

#### Study governance and quality control

The study field staff all underwent Good Clinical Practice training in addition to formal training in all study activities as outlined in the Standard Operating Procedures prior to study initiation. This included formal training in study information, informed consent, survey administration, electronic data capture, biological sample collection and transportation. Intermittent spot checks will be carried out of field data collection activities, and regular team meetings will be held to facilitate discussion of study progress, challenges in the field, and for performance-based feedback based on review of incoming study data. Photographs of a random 10% sample of urine dipsticks will be reviewed by the investigator team. In the laboratory, calibrators and controls will be used to ensure reliability and consistency of assay results; in addition to use of standardized operating procedures for sample processing (see ‘Laboratory Methods, Quality control’). Spot checks will be carried out to ensure the temperature of the cool boxes used for sample transportation are maintained within the required range.

### Patient and public involvement

Prior to initiation of this study, approval for the study was granted by the Karonga District Health Office, and meetings were held with community leaders through the Area Development Committee. HDSS residents had been engaged at an individual- and community-levels on all aspects of the wider MEIRU programme of census activities, HLM LTC survey and its nested studies, including this study, through a series of open community meetings to raise awareness and provide opportunities for questions and discussion. Additional meetings with group village heads and community representatives of each area of the HDSS due to be specifically visited by this study will take place one to two weeks before initiation of the study in that particular area, to discuss finer details of this study, enabling wider dissemination of information by community representatives and providing a more focused opportunity for questions and concerns to be raised ad addressed.

On completion of the study, community meetings will be held to feedback the results of the study and wider HLM-LTC survey, including with the more recently formed community advisory group. This will include opportunity for questions, discussion, and priorities to be raised with regards to future research directions.

## Discussion

The methodology of the Impso study, an ongoing prospective population cohort study aiming to determine risk factors for progressively impaired kidney function amongst adults living in rural northern Malawi, is described.

This study has been designed as a prospective cohort study nested within two large, rural population-based surveys of long-term health conditions, in order to leverage existing longitudinal follow-up data, including stored paired, individual-level serum samples. The overall study’s sample size is limited by the available resources (time and funds). This has necessitated convenience sampling of individuals within the HDSS with pre-existing baseline eGFR results available, calculated from serum cystatin C and creatinine already tested on serum samples collected in previous studies. In order to ensure the internal and external validity of the study findings, the characteristics of individuals who participate in the current study will be compared to those of the general population of the HDSS. We have already compared the characteristics of individuals with and without availability of existing cystatin C test results within the wider HDSS adult population **[Supplementary file S3].**

We are targeting recruitment to individuals with a baseline eGFR_cysc_ less than 90 ml/min/1.73m^2^ in order to prioritise inclusion of individuals with existing evidence of at least mildly impaired kidney function. It is known from other settings that the risks of adverse clinical outcomes including incident kidney failure and mortality are highest in individuals with lower baseline eGFR, specifically those with a baseline eGFR less than 60 ml/min/1.73m^2^, proteinuria, and history of prior rapid eGFR decline, amongst other clinical risk factors.(70–73) In some settings, studies of older adults and people living with HIV, have also suggested an increased risk of CKD development among individuals with baseline eGFR between 60 and 90 ml/min/1.73m^2^ compared to eGFR ≥ 90 ml/min/1.73m^2^.(74, 75) It is not yet known if the clinical cut-offs used to define normal range for eGFR (≥ 60ml/min/1.73m^2^) and CKD classification are appropriate in African settings.(13, 76) Targeting inclusion to individuals with eGFR_cysc_ < 90 ml/min/1.73m^2^ will allow us to interrogate in more detail the differences in clinical outcomes between individuals with baseline eGFR either side of established classification values. Inclusion of individuals with eGFR < 90 ml/min/1.73m^2^ is also of more clinical relevance as most laboratories do not routinely report actual eGFR values that are ≥ 90 ml/min/1.73m^2^.(77, 78)

It is important to note that in some study populations, higher baseline eGFR attributed to hyperfiltration can be associated with rapid eGFR decline(79–82). We may therefore fail to capture some of these high-risk individuals in our study. However, this phenomenon is mostly observed in specific conditions such as diabetes and metabolic syndrome, and not necessarily in other patient groups(83, 84). We have already compared the baseline characteristics of individuals with baseline eGFR_cysc_ < 90 ml/min/1.73m^2^ and (≥ 90ml/min/1.73m^2^). **[Supplementary file S4].** To build understanding of longer-term health of those not recruited, we plan to investigate differences in clinical characteristics, eGFR measures and all-cause mortality (to be reported in a separate survival analysis) between individuals recruited and not recruited to the study. We will be adjusting for baseline eGFR in our analysis. Furthermore, although our recruitment criteria are based on baseline eGFR_cysc_ criteria, we also plan to look at a range of outcome measures using eGFR_creat_, meaning that we will have outcome data for individuals with higher starting eGFR within the eGFR_creat_ analysis, and will also be able to interrogate outcome measures according to baseline eGFRdiff)(85, 86).

Overall the inclusion criteria for this study are broad, including measures to facilitate inclusion of adults with reduced mental capacity to provide consent, for example due to mental health conditions, learning disability and dementia; often neglected groups in clinical research.(87) Exclusion of individuals pregnant at the time of either historical (baseline) blood sampling or at the time of the current study is due to the known temporary physiological changes in eGFR that occur during pregnancy. Inclusion of these individuals would limit conclusions about overall longitudinal trends in eGFR using on the available blood samples collected at these time points. For individuals hospitalised for over 24 hours, biological sample collection will be delayed until three months post-discharge due to the known high prevalence of acute kidney injury among hospital inpatients, including in low-resource settings – often undiagnosed.(88)

Strengths of this study include the relatively long duration of follow-up (anticipated to be over nine years on average), and richness of paired, individual-level participant risk factor data from linked studies including sociodemographic, anthropometric, clinical and biochemical data. We anticipate a high (>85%) participation rate in line with that seen in other studies conducted in the Karonga HDSS.(52) A particular strength of this study is that it is the first study use of both cystatin C- and creatinine-based eGFR to examine predictors for longitudinal decline in kidney function in an African setting. This is important because creatinine-based estimates significantly underestimate the burden of CKD in African settings, including in Malawi, when compared to Iohexol plasma clearance mGFR; in contrast, cystatin C based eGFR more closely mirrors mGFR.(13) Use of both measures will therefore enable more accurate determination of longitudinal trends in eGFR decline, and association with different risk factors, in this Malawian setting, and to compare findings using the two methods for estimating GFR.

This study also has limitations. Being conducted in rural Karonga, findings may not be generalisable to other regions of Malawi, or to other African settings, in particular urban areas, although over 80% of Malawi’s population live in rural areas.(89) In addition, a maximum of three longitudinal eGFR measures per participant will be available, limiting the ability to precisely determine annual rate of eGFR decline. However, we will be able to assess a range internationally-recognised outcome measures for sustained eGFR decline and CKD progression, as well as new development of CKD, due to the fact that the two most recent measures for each participant will be collected more than three months apart. This is in contrast to many cross-sectional studies of CKD prevalence conducted in sub-Saharan Africa, which have commonly had to rely on a single eGFR measures, limiting ability to distinguish between acute and chronic kidney disease.(8–10)

This study will be limited by survivorship bias, as individuals who are older and those who have more advanced kidney disease at baseline, including those who have already progressed to untreated kidney failure, are more likely to have died during the years preceding this study. The CKD risk factors we can interrogate will be limited by availability of measurements and information that can be collected without limitation by significant information bias. Pregnancy status will be based on self-report rather than testing so it is possible that some individuals in early pregnancy may be recruited into the study. Furthermore, as this is set in an open population cohort, some potentially eligible individuals may have left the area. The fact that rich baseline data are available for all individuals will allow interrogation of differences between individuals who do, and do not, participate or meet eligibility for the study.

The sample size for this study is relatively small, albeit adequate to detect reasonable effect size estimates (OR 1.66 – 4.38) for the association of different risk factors with progressive decline in kidney function. There is limited power to detect more modest effects, as well as the effects of less common risk factors. Our outcome measures for CKD progression are guided by those recommended by KDIGO however it is important to note that these were not derived from African populations.

Finally, the use of the Jaffe colorimetric kinetic method for quantification of serum creatinine, with its inferior accuracy to the recommended enzymatic method(29, 90, 91) is a limitation. The decision to use this method is based on the fact that the individual paired serum samples for study participants collected in the earlier linked studies have already been tested using the Jaffe method – as occurs in over 90% of studies from Africa due to resource limitations.(10) In order to determine eGFR change over time, the same quantification method must therefore be used to test the newly collected serum samples in this study.(92) In mitigation, the primary outcome measure of this study will use eGFR_cysc_ rather than eGFR_creat._ Furthermore, serum samples will be centrifuged within a maximum of eight hours from the time of collection; well within the recommended 24 hour window after which delay in sample separation can add significant positive bias when using the Jaffe method.(93, 94) The consent obtained for long-term storage of all serum samples will enable testing and analyses using the enzymatic method for creatinine quantification to be conducted as part of future work.

In conclusion, the Impso study will be one of very few longitudinal studies to date to study risk factors for progressive decline in kidney function in a low-resource, rural, population-based African setting, and the first in this setting to use the more sensitive kidney function biomarker, cystatin C, to examine longitudinal eGFR trajectories. This study is likely to contribute important data about individuals at the highest risk of rapid eGFR decline and CKD progression in rural Malawi, markers for multiple adverse health outcomes.(70, 73) This will inform future research into specific risk factor exposures of importance, and could directly inform future health policies in Malawi for targeting CKD screening, prevention and treatment strategies to the highest risk patient groups.

## Data Availability

All relevant data are within the manuscript and its Supporting Information files.

## Acknowledgements

CS is also affiliated with Malawi Epidemiology and Intervention Research Unit (MEIRU) and with the Malawi Liverpool Wellcome Research Programme, Blantyre, Malawi.

We thank participants of all the named studies in addition to the wider MEIRU research, management and support teams for their contribution. We thank Professor Segun Fatumo (Queen Mary, University of London) for his support in provision of serum creatinine and cystatin C assays. We acknowledge the wider ARK consortium for their work pre-dating this study and ongoing collaboration. CS also thanks the Liverpool Clinical PhD Programme for Health Priorities in the Global South (Professor Neil French, Elly Wallis, and steering group) for their support and the Liverpool School of Tropical Medicine for sponsorship of this study.

## Author contributions

The current study was conceived and designed by CMS, AJP, ACC, HCM, DMT, FL, JF and RK. The study builds directly upon previous linked research studies which were conceived and designed by AJP, ACC, WN, JF and RK. CMS, AJP, FL, and ACC applied for and acquired funding for the study. Surveys were designed by CMS, AJP and DMT; with programming of data collection tools implemented and managed by PK, BBM and DN with support from the wider MEIRU data team. Study standard operating procedures were written by CMS and CM with input and revisions from SMM, PK, AJP, AD, CP and DMT. Community sensitisation work was planned by CMS, AD, FK and conducted by CMS, AD, FK, LK and DB. Staff training was conducted by CMS, AD, TNG, SMM and CM. Pilot data was collected by LK and DB, with supervision by CMS, AD and TNG. Data management systems for the pilot data were managed by PK and BBM. Laboratory procedures were supervised by CM. The manuscript was drafted by CMS. All authors contributed to critical review and revisions of the manuscript prior to publication apart from LK^†^. This work was conducted as part of CMS’ PhD fellowship; CMS is supervised by AJP, FL, DMT and HCM with collaboration from ACC, JF and RK.

## CRediT authorship contribution statement

**Charlotte M Snead:** Conceptualization, Methodology, Formal Analysis, Resources, Data Curation, Supervision, Investigation, Validation, Formal Analysis, Project Administration, Funding Acquisition, Writing – Original Draft, Writing – Review and Editing, Visualization. **Chimwemwe Mkandawire**: Methodology, Investigation, Validation, Resources, Supervision, Project Administration, Writing – Review and Editing. **Paul Kambiya**: Software, Validation, Resources, Data Curation, Project Administration, Writing – Review and Editing. **Shekinah Munthali-Mkandawire**: Methodology, Supervision, Project Administration, Writing – Review and Editing. **Fredrick Kalobekamo**: Investigation, Supervision, Project Administration, Writing – Review and Editing. **Albert Dube:** Methodology, Supervision, Project Administration, Writing – Review and Editing. **Thandile Nkosi-Gondwe**: Supervision; Project Administration; Writing – Review and Editing. **Baltazar Bananga Mtenga:** Software, Validation, Resources, Data Curation, Writing – Review and Editing. **Dominic Nzundah**: Software, Data Curation, Writing – Review and Editing**. Lenford Kwamkwanya**: Conceptualization, Investigation. **Desire Bellings**: Conceptualization, Investigation, Writing – Review and Editing**. Wisdom Nakanga**: Conceptualization, Methodology, Project Administration, Writing – Review and Editing. **June Fabian:** Conceptualization, Methodology, Resources, Supervision, Formal Analysis, Writing – Review and Editing. **Robert Kalyesubula:** Conceptualization, Methodology, Resources, Supervision, Formal Analysis, Writing – Review and Editing. **Chimota Phiri**: Methodology, Supervision, Writing – Review and Editing. **Felix Limbani:** Conceptualization, Methodology, Supervision, Writing – Review and Editing. **Dominic M Taylor:** Conceptualization, Methodology, Supervision, Writing – Review and Editing. **Amelia C Crampin:** Conceptualization, Methodology, Resources, Supervision, Funding Acquisition, Formal Analysis, Writing – Review and Editing. **Henry Mwandumba:** Conceptualization, Methodology, Supervision, Writing – Review and Editing. **Alison J Price:** Conceptualization, Methodology, Supervision, Funding Acquisition, Writing – Review and Editing.

## Footnotes

1 In the event of internal migration within the HDSS, resident status is maintained.

2 Commencement of maintenance KRT forms part of the KDIGO definition of kidney failure, however we do not expect to find any individuals meeting this outcome criterion in our cohort.

